# 2-mercaptoethanol (2-ME)-based IATs or polybrene method mitigates the interference of anti-CD38 immunotherapy on blood compatibility tests

**DOI:** 10.1101/2020.12.14.20238311

**Authors:** Ye Zhou, Leiyu Chen, Tianshu Jiang, Liangfeng Fan, Hang Lei, Yuqing Wang, Hasiyati Heililahong, Jianqing Mi, Danxin Du, Tianhong Miao, Rong Xia, Xuefeng Wang, Dong Xiang, Xiaohong Cai, Xiaofeng Tang

**Affiliations:** Department of Blood Transfusion, Changzheng Hospital, Naval Medical University, Shanghai, China; Blood Group Reference Laboratory, Shanghai Blood Center, Shanghai, China; Department of Blood Transfusion, Laboratory Diagnosis Center, Ruijin Hospital, School of Medicine, Shanghai Jiaotong University, Shanghai, China; Department of Hematology, Ruijin Hospital, School of Medicine, Shanghai Jiaotong University, Shanghai, China; Department of Blood Transfusion, Huashan Hospital Fudan University, Shanghai, China; Blood Group Reference Laboratory, Beijing Red Cross Blood Center, Beijing, China

**Keywords:** Daratumumab, anti-CD38, 2-mercaptoethanol, Polybrene, blood transfusion

## Abstract

**Background:** Treating red blood cells (RBCs) with dithiothreitol (DTT) is a wildly-recommended to overcome the interference of anti-CD38 immunotherapy with blood compatibility testing. Nevertheless, DTT can be hard to obtain in clinical laboratory, while its use in routine practice may be time-consuming. In the following study, we explored the feasibility of using a commercial 2-mercaptoethanol (2-ME) working solution or the time-saving polybrene method to mitigate the daratumumab (DARA) interference.

**Materials and Methods:** Antibody screening and cross-matching were performed using 2-ME or DTT-based indirect antiglobulin tests (IATs) and polybrene method (human IgG anti-E same IATs titer as DARA as positive control) on 37 samples, and these samples were from patients enrolled in the “Several methods resolve the interference of anti-CD38 monoclonal antibody on blood compatibility tests” clinical trial (www.chictr.org.cn identifier: ChiCTR2000040761). Most clinically important blood group antigens on RBCs were detected after treatment with 2-ME or DTT. Hemoglobin values were compared after 69 units RBCs were transfused with compatible cross-matching results by a 2-ME-based method or polybrene test.

**Results:** Treating RBCs with 2-ME eliminates the DARA interference with the antibody screening or cross-matching; yet, K antigen is denatured during treatment. DARA with 2+ agglutinations of anti-E control does not interfere with antibody screening and cross-matching via polybrene method. After RBCs transfusion with a negative cross-matching test by 2-ME-based IATs or polybrene method, hemoglobin significantly increased without adverse transfusion reactions.

**Conclusion:** 2-ME-based IATs or polybrene method could be used to mitigate DARA interference as DTT.

## Introduction

CD38 molecule is a multifunctional type II transmembrane glycoprotein that participates in cell adhesion and transmembrane signaling process^1^. CD38 is highly expressed in hematological malignancies, including multiple myeloma (MM); thus, it is an ideal target for immunotherapy.

Daratumumab (DARA) is the first immunoglobulin G1κ (IgG1κ) full-human CD38 monoclonal antibody that specifically binds to CD38 antigen epitopes expressed on the surface of tumor cells^2^. DARA has been primarily used to treat patients with MM. Daratumumab targets CD38 on myeloma and facilitates antibody-dependent cell-mediated cytotoxicity (ADCC), antibody-dependent cellular phagocytosis (ADCP), complement-dependent cytotoxicity (CDC), and antibody cross-linking mediated apoptosis and indirect immunity^3^. However, considering that CD38 is also expressed in red blood cells (RBCs), although at a lower level compared to malignant cells, the drug may also bind to healthy cells producing high blood pan-reactivity and incompatibility ^4-6^

It has been found that significant para-agglutination reactions occur after the clinical use of DARA^7,8^. DARA’s presence in the sera can interfere with pretransfusion testing; thus, handling samples from patients treated with this drug requires a special strategy. If the history of CD38 monoclonal antibody use in clinic is not recorded, the presence of accidental antibodies in RBCs of patients may be mistakenly interpreted, thus leading to false positives and interfering with blood matching, as well as difficulties in clinical blood transfusion. On the contrary, in some cases, false-positive may mask the presence of clinically significant alloantibodies due to CD38 antibody interference, thus increasing the risk of delayed hemolytic transfusion reactions.

Currently, several methods, including the application of sulfhydryl reagents (e.g., dithiothreitol, DTT), polybrene and proteolytic enzymes (trypsin/papain), RBC phenotyping, RBC genotyping, and neutralizing proteins (anti-antibodies and soluble CD38 protein) have been reported to overcome the interference of DARA on transfusion compatibility testing. Treatment of RBCs with DTT is the most common method to remove DARA interference. As a thiol reducing agent, DTT can cleave disulfide bonds in the extracellular region of CD38 molecules so as to denature CD38 antigen and prevent it from binding to the CD38 antibody. Treatment of RBCs with DTT provides a safe blood supply for patients receiving CD38 antibody therapy^9^. However, in some countries such as China, DTT is not conventionally used in clinic. On the other hand, 2-mercaptoethanol (2-ME), the same sulfhydryl reducing agent, is routinely used and easily available in China. Chapuy *et al* suggested that the use of 2-ME might remove the interference induced by DARA^10^; yet, it has not been confirmed by experiments. It was reported that the Polybrene method avoided interference of DARA on the compatibility of blood transfusion^11,12^; however, there is still a lack of large-sample, multi-center research and demonstration.

In this study, we treated RBCs of blood donors with 2-ME so as to avoid the interference of DARA. Meanwhile, we performed a transfusion compatibility test with the Polybrene method in 24 patients with MM using DARA and tracked the clinical outcomes after transfusion.

## Materials and Methods

### Patients

A total of 24 DARA-treated MM patients were involved in this study, who were enrolled in the “Several methods resolve the interference of anti-CD38 monoclonal antibody on blood compatibility tests” clinical trial (www.chictr.org.cn identifier: ChiCTR2000040761). Among these patients, 12 patients were admitted to Changzheng Hospital, 5 to Ruijin Hospital and 7 patients’ samples were sent to the blood group reference laboratory of Shanghai Blood Center between March 2018 and April 2020. All patients received one or more than one blood transfusion (total 37 times).

### Tests before the first transfusion of DARA-received patient samples

Antibody screening tests for MM patients receiving DARA were performed with an IH-1000 automatic blood grouping analyzer (BIO-RAD, Roanne, France) or ORTHO Vision Max analyzer (Ortho, Mannedorf, Switzerland) for ABO, RhD blood group, and IATs (indirect antiglobulin tests) detection.

### DTT or 2-ME treatment of RBCs

Two different brands of 2-ME working solution (Baso, Zhuhai, China; SHPBC, Shanghai, China) with clinical testing qualifications were used for RBCs, including antibody screening cells (SHPBC, Shanghai), panel RBCs (Sanquin, Amsterdam, Netherlands), single RBCs from blood donors. When treating donor RBCs with 2-ME, DTT (Roche, Mannheim, Germany) was also used to treat the same cells, in order to compare the treatment effects between these two reagents. The DTT treatment of RBCs was carried out following the AABB technical manual^13^. The 2-ME treatment of antibody screening RBCs, antibody identification panel cells, or donor RBCs was performed as follows: a total of 200μl of 3% RBC solution were washed four times with PBS (pH 7.4) before adding 800μl of 2-ME in each tube. The mixture was incubated at 37°C for 30 minutes, mixing by inversion every 7 to 8 minutes. The cells were then washed four times with PBS (pH 7.4) and re-suspended in 200μl of LISS (Low Ionic Strength Solution).

### IATs or cross-matching of DARA-received patient samples using DTT or 2-ME treatment RBCs

Antibody screening cells treated with 2-ME were used for antibody screening. Donor RBCs treated with 2-ME were used for cross-matching by microcolumn gel method using Polyspecific Anti-Human Globulin Anti-IgG,-C3d Card (BIO-RAD, Cressier, Switzerland). RBCs positive for the E and K antigens were used as positive and negative controls to verify 2-ME treatment efficacy.

### Effect of 2-ME treatment on RBC blood group antigens

Anti-K, anti-D, anti-M, anti-N, anti-S, anti-s, anti-Jka, anti-Jkb, anti-Fya, anti-Fyb, anti-Dia IgG antibody reagents were purchased from CE-immundiagnostika GmbH, Germany. Anti-E, anti-e, anti-C, anti-C IgG antibodies were in-house human IgG antibodies. Anti-K, anti-D, anti-E, anti-e, anti-C, anti-c, anti-M, anti-N, anti-S, anti-s, anti-Jka, anti-Jkb, anti-Fya, anti-Fyb, anti-Dia antibody reagents were diluted with donor anti-screening negative AB plasma to achieve titer 1 (polyspecific anti-human globulin anti-IgG, -C3d card (BIO-RAD, Cressier, Switzerland)). Antibody identification panel RBCs treated with 2-ME containing corresponding antigens reacted with diluted corresponding antibody reagents in polyspecific anti-human globulin cards. The manufacture instructions were followed to observe whether 2-ME had a destructive effect on RBC antigens.

### IATs or cross-matching of DARA-received patient samples using polybrene method

In order to verify the detectability of polybrene method for clinically significant antibody, antibody titers were standardized between plasma of DARA-treated patients and ABO compatible human IgG anti-E plasma by IATs using polyspecific anti-human globulin anti-IgG, -C3d card. Anti-E plasma was diluted by AB-type plasma. The plasma (negative for RBC blood group antibody screening) from patients receiving DARA treatment or diluted series of anti-E plasma reacted with RBC suspension from the same E-positive donor in anti-human globulin cards. The anti-E control plasma dilution, whose agglutination intensity was consistent with the patient’s, was selected as the control after dilution of anti-E plasma for subsequent antibody screening and cross-matching by polybrene method. The polybrene kit was purchased from SHPBC (Shanghai, China) and the manufacture instructions were followed. The cross-matching donor cells were E-positive RBCs, and the results were synchronously compared with the previously standardized human equivalent IgG anti-E plasma as the control.

### Evaluation of efficacy of blood transfusion in MM patients treated with DRAR

69 RBC units negative for cross-matching by 2-ME and polybrene method was transfused to 24 patients and the Hb values were detected before and after transfusion. Blood transfusion efficacy was evaluated with a two-sided paired t-test using GraphPad Prime (GraphPad Prism, Version 8.0, GraphPad Software, Inc., San Diego, CA).

## Results

### Characteristics of routine pre-transfusion compatibility testing in Chinese patients undergoing DARA treatment

During the routine pre-transfusion detection of 24 Chinese patients treated with DARA, ABO, RhD, and other Rh phenotyping tests were not affected. Antibody screening (**Fig. 1A**, left) or cross-matching (**Fig. 1B**, left) was positive. The agglutination intensity was usually approximately 2+.

**Fig.1.**
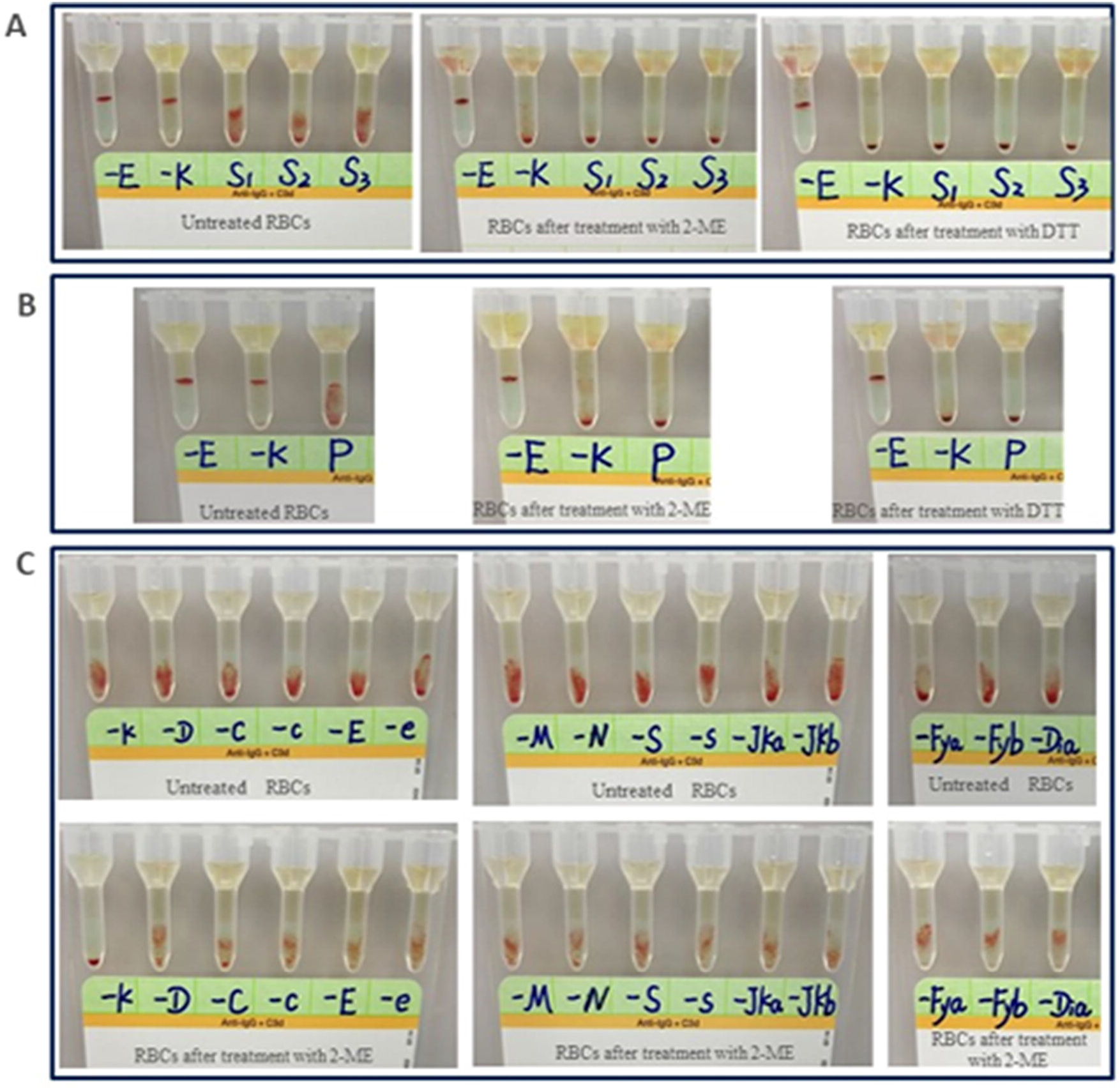
Pre-transfusion compatibility test using IATs in Chinese patients undergoing DARA treatment. **(A)** Antibody screening results of patients before and after 2-ME/DTT treatment. **(B)** Cross-matching results of patients before and after 2-ME/DTT treatment. **(C)** Antigen detection results before and after 2-ME treatment in panel RBCs.

### Antibody screening and cross-matching results of RBCs treated with 2-ME

The antibody screening test based on the microcolumn gel method was performed on the antibody screening RBCs treated with 2-ME and plasma of MM patients after receiving DARA. The result was negative (**Fig. 1A**). When the donor RBCs treated with 2-ME were cross-matched using a microcolumn gel method, pan-reactivity previously detected in the plasma of these patients by IATs was not observed (**Fig. 1B**). The efficacy of treatment was proved by the denaturation of the K antigen in the K+ control cell and preservation of the E antigen in the E+ control cell. The experimental results are consistent with those of DTT treatment of RBCs **(Fig. 1A**, right; **Fig. 1B**, right).

### Effect of 2-ME treatment on RBC blood group antigens

Serious hemolysis was observed when reagent RBCs were treated with 2-ME (SHPBC) at 1:4 ratio, while no obvious hemolysis was seen until the end of incubation when reagent RBCs were treated with 2-ME (Baso) at 1:4 ratio. The antibody identification panel cells treated with the latter reacted with the corresponding antibody reagents in the anti-human globulin card. 2-ME had a destructive effect on K antigen, but not on other RBC antigens (**Fig. 1C**).

### Antibody screening and cross-matching tests of MM patients treated with DARA using Polybrene method

The agglutination intensity of human IgG anti-E plasma detected by gel card at different dilutions is shown in **Fig. 2A**. For each sample, human IgG anti-E was standardized to a dilution of 128-256 to show the same agglutination intensity as the patient’s CD38 monoclonal antibody in IATs. The results of both antibody screening (**Fig. 2B**) and cross-matching **(Fig. 2C**) using Polybrene method were negative, while the result of IgG anti-E plasma with a titer of 128-256 was positive for 2+ agglutination intensity.

**Fig. 2.**
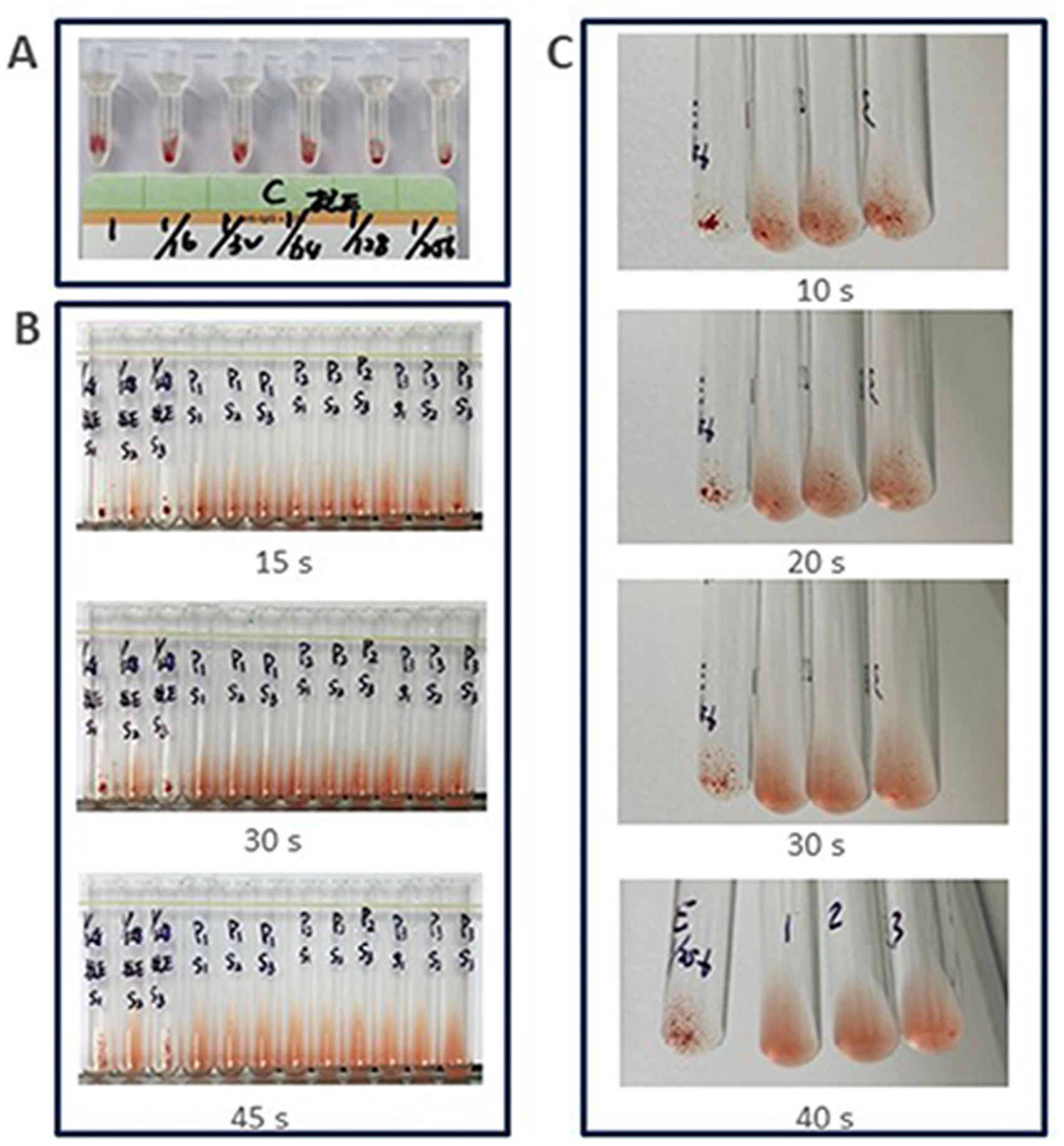
Pre-transfusion compatibility test using polybrene method in Chinese patients treated with DARA. The results were all negative, while IgG anti-E plasma showed a 2+ agglutination intensity. **(A)** 1:16-1:256 agglutination intensity of human IgG Anti-E in IATs at different dilutions. **(B)** Antibody screening (from left per three tubes: anti-E control, Patient 1(P1), Patient 2(P2), Patient 3(P3);S1-S3 means 3 antibody screening cells). **(C)** Major cross-matching results (from left: anti-E control, Patient 1, 2, 3)

### Transfusion efficacy in patients with MM treated with DARA

RBC units of IATs cross-matching negative using donor RBCs treated with 2-ME or Polybrene cross-matching negative were infused into 24 patients with MM after DARA. No adverse transfusion reactions were observed in clinic. For each 1 unit RBCs transfusion per average, Hb was significantly increased compared with that before transfusion (**Fig. 3**).

**Fig. 3.**
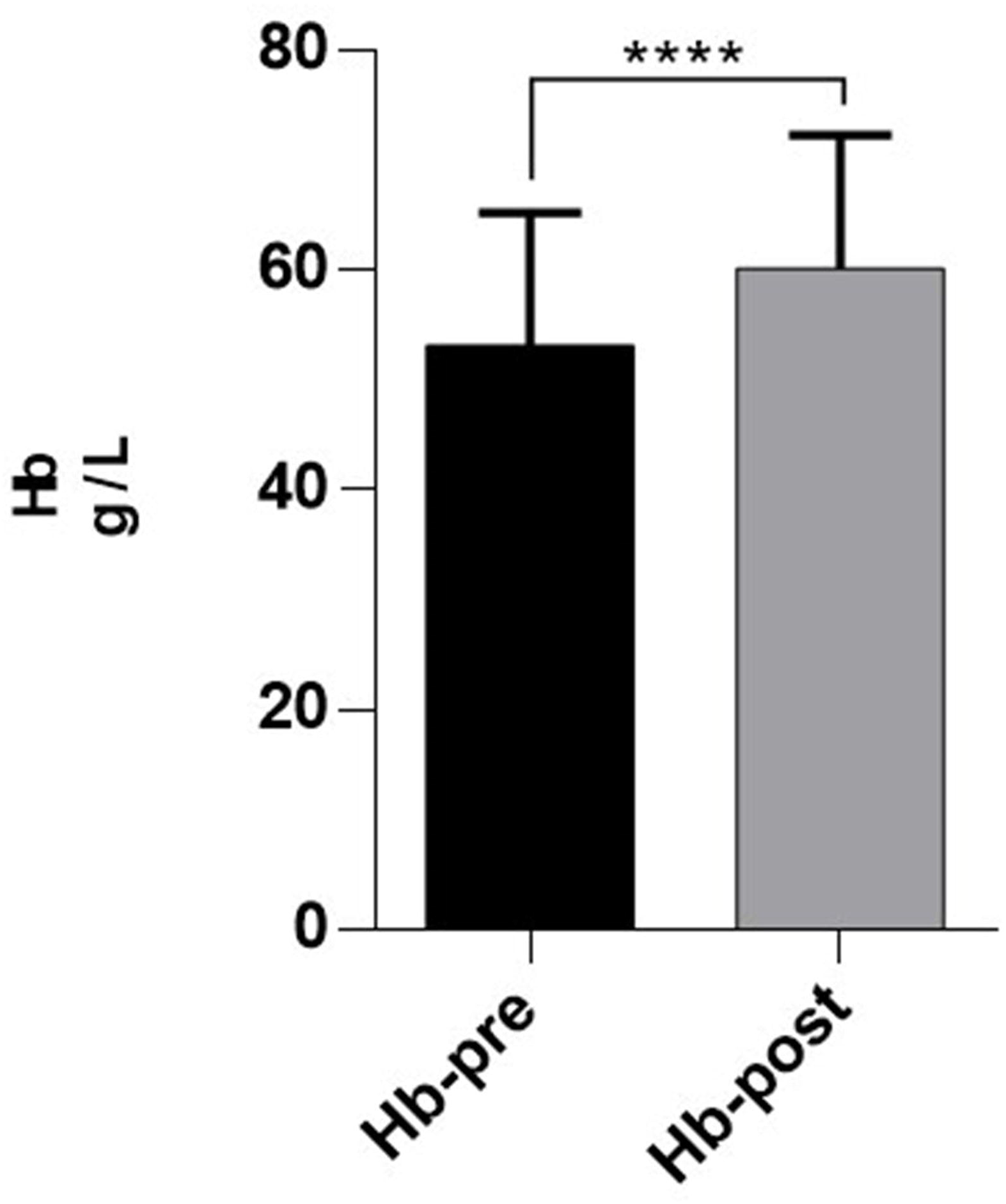
Transfusion efficacy in MM patients treated with DARA. There is a significant difference in patients Hb values before and after infusion (P<0.0001). Hb is significantly increased in patients with 1 unit RBCs transfusion.

## Discussion

Considering that CD38 is expressed on RBCs, it may cause RBC agglutination after the addition of anti-globulin reagents^14^. Interference with agglutination reactions can be observed regardless of the type of detection (antibody screening, antibody identification, IAT, DAT, Crossmatching test) based on anti-globulin reagents and media (e.g., gel, tube, solid phase)^7, 9, 15^. The reaction can continue for six months after discontinuation of DARA^7,8^, which is consistent with our results of Chinese patients. Only tube tests, such as ABO and RhD, as well as other Rh phenotyping tests, are not disturbed, which indicates that weak expression of CD38 on RBCs generally does not produce agglutination in saline medium. Previous studies have shown that DTT treatment of RBCs can remove DARA interference on the pre-transfusion compatibility test. In addition, it has been speculated that 2-ME can have a similar effect as DTT ^10^. Our study demonstrated that some commercial types of 2-ME can achieve the same treatment effect as DTT, either in antibody screening or cross-matching tests, though certain brand have been associated with severe hemolysis. In China, DTT hasn’t clinical testing qualification but 2-ME has. A 2-ME working solution can be directly applied with less relative toxicity, thus, it is more suitable for clinical use.

Thiol reductants can denature other important RBC antigens, such as Kell system antigens^14^. In this study, we found that the 2-ME antigen destroys the Kell system but has no obvious destructive effect on other blood group system antigens, which is similar to DTT. Kell system has polymorphism distribution in the Caucasian population. Thus, Kell homotypic transfusion is recommended for Caucasian patients in need of a blood transfusion after using DARA so as to compensate for the failure of antigen-antibody reaction in this system due to the destruction of the Kell system antigen after DTT treatment of RBCs. The East Asian populations usually carry a k+K-type. The K+blood group has a frequency of about 0.07%, thus, Kell blood grouping is not commonly performed^16^. Consequently, Kell blood grouping after sulfhydryl reductant treatment of RBCs is not necessary for the Asian population, including the Chinese.

Similarly, the widespread use of the polybrene method in pre-transfusion testing in the East Asian population is based on this consideration. Polybrene is a rapid and straightforward method for RBC antibody detection, which is used for pre-transfusion antibody detection and cross-matching test in many clinical laboratories across Southeast Asian countries. The result of anti-globulin testing after using DARA is positive, while that of polyamine test is negative. At present, the use of polybrene method to overcome the interference of DARA on pre-transfusion compatibility detection is scattered in few reports^11,12^. The results of 37 pre-transfusion compatibility tests in this study confirmed that the Polybrene results did not interfere with the CD38 monoclonal antibody.

We speculate that the underlying mechanism is similar to the polybrene method’s weak detection ability for Kell blood group antigens. Kell protein, like CD38 molecule, is a type II transmembrane glycoprotein with a single transmembrane, and a carboxyl group outside the cell. Polybrene is a small molecular weight quaternary ammonium that binds to negatively charged molecules on the surface of RBCs^17^. Polybrene method has a weak detection ability for the Kell system’s antibody, which is related to the negatively charged carboxyl group at the Kell protein terminal. The positively charged polybrene molecule binding to the negatively charged Kell protein molecule in a low ionic medium environment interferes with the Kell system’s antigen-antibody reaction. Based on the same mechanism, the polybrene test is insensitive to CD38-related antigen-antibody reaction. Under a low ionic environment, the CD38 molecule carries a negatively charged carboxyl group extracellularly, which easily binds to the polybrene molecule and interferes with the binding of the CD38 molecule to DARA.

In this study, although the patient results of both antibody screening and cross-matching using Polybrene method were negative, human IgG anti-E antibody which displayed the same agglutination intensity as DARA in IATs, was 2+ positive. This indicates that antibody screening and cross-matching with Polybrene method would not miss human IgG blood group antibodies. Since there is no other method to remove CD38 molecules from RBCs without destroying the Kell system’s antigens, polybrene method can also be used for pre-transfusion testing of Caucasian people under the premise of combining Kell homotypic transfusion. However, large-sample studies are needed to further verify these findings.

After RBCs transfusion with a negative cross-matching test by 2-ME-based IATs or polybrene method, all the patients showed a significant increase in Hb (according to the average amount of 1 unit per infusion). No adverse transfusion reactions were observed in the clinic, indicating the safe use of these methods to remove DARA interference on the pre-transfusion compatibility test.

In conclusion, our data suggest that 2-ME treatment on RBCs may have a similar effect as the DTT in patients with MM who received DARA treatment. 2-ME can remove the interference of DARA with pre-transfusion testing. However, a large amount of hemolysis and other obvious abnormalities of RBCs during the treatment should be avoided. Polybrene method can be used for these patients for cross-matching or antibody screening. Still, weak positive control should be applied to prevent the missed detection of clinically significant weak antibodies. When using the thiol reagent or polyamine method, the Kell blood group system’s polymorphism should be combined in different races to determine whether the homotypic transfusion of this blood group is synchronously performed.

## Data Availability

The data used to support the findings of this study are available from the corresponding author upon request

## Statement of Ethics

The study was approved by the ethics committees of Changzheng Hospital,Shanghai Blood Center, Ruijin Hospital, Huashan Hospital, Beijing Red Cross Blood Center. Written informed consent was obtained for each patient.

